# Who infected the reported cases? Evidence from 678,482 COVID-19 cases with identified infector collected in routine surveillance in the Netherlands, 2020-2022

**DOI:** 10.64898/2026.05.15.26347859

**Authors:** Jantien A. Backer, Ka Yin Leung, Stijn P. Andeweg, Jan van de Kassteele, Irene Veldhuijzen, Susan Hahné, Jacco Wallinga

**Affiliations:** Centre for Infectious Disease Control, National Institute for Public Health and the Environment (RIVM), Bilthoven, the Netherlands; Biomedical Data Sciences, Leiden University Medical Center, Leiden, the Netherlands

## Abstract

**Background:** During infectious disease outbreaks, characteristics of reported cases are routinely collected. These give information on becoming infected but not on infecting others. We assess whether linking infectees to infectors, together with their characteristics, can help understand transmission.

**Methods:** From the start of the COVID-19 pandemic in the Netherlands, reported cases were asked to identify their most probable infector in routine surveillance, enabling the linking of cases. We assess for the period 27 February 2020 – 11 April 2022 whether the infectees of these transmission pairs are representative of all reported cases, whether the transmission pairs yield verifiable estimates of epidemiological characteristics (here the serial interval), and whether they provide additional information on transmission.

**Results:** Of 8,003,008 reported cases, 678,482 (8.5%) could be linked to their most probable infector. These infectees were largely representative of the reported cases regarding age group, sex, and geographical location. The mean serial interval of 3.6 days (sd 3.4 days) from transmission pairs aligns with literature. Transmissions between age groups largely follow known contact patterns. Most transmissions in September 2021 occurred between persons who were not (fully) vaccinated, indicating the effectiveness of the vaccine, and relatively few between persons with different vaccination status, suggesting assortative mixing in vaccination status.

**Conclusion:** Transmission pairs can be efficiently collected in routine surveillance, providing insight into disease transmission. The current post-pandemic period provides an excellent opportunity to adjust reporting systems for linking infectees to their most probable infector as preparation for future outbreaks.

## Introduction

When a completely new or new variant of an infectious disease pathogen emerges, its characteristics are initially unknown. Basic epidemiological variables, such as the incubation period or age-specific transmission risks, are needed to inform an appropriate public health response. Knowledge about the transmission dynamics can help design targeted interventions to mitigate the outbreak as effectively as possible.

Studies that determine who infected whom are invaluable to gain insight into the transmission dynamics. Detailed reports of transmission events at the start of an epidemic can provide the first estimates of epidemiological characteristics, as was shown for COVID-19 in 2019 [1–4] and Mpox in 2022 [5]. Information on infectees and their infectors can also be obtained from household studies where all household members are monitored over time, from which for instance age-dependent transmission characteristics for influenza [6,7] and COVID-19 [8–10] were obtained. Another data source from which transmission pairs can be constructed is contact tracing, where contacts of reported cases are followed over time, as was done for COVID-19 [11,12] and pandemic influenza in 2009 [13]. Genomic sequencing can provide additional evidence as to whether viral genomes of an infector and infectee are sufficiently similar to support a transmission link without inferring its direction. These types of studies have in common that they require time-intensive and costly collection methods. As a result, they typically provide reliable data, but only for a limited number of cases and over a limited time span.

An alternative method to collect transmission pairs was introduced at the start of the COVID-19 pandemic in the Netherlands. In the routine surveillance questionnaire that was filled out for all reported cases [14], a question was included to identify the most probable infector. If the indicated infector was also a reported case in the surveillance system, the two cases and all their characteristics, such as age, location and symptom onset date, were linked as a transmission pair. The identification of the infector relied on the assessment of the infectee with the help of a public health officer and was not confirmed by genomic sequencing or other methods. Transmission links were available in large numbers over the entire period of the COVID-19 pandemic, without substantial additional costs to the routine surveillance system.

These routinely collected transmission pairs were used to study the role of children in SARS-CoV-2 transmission [15], the interaction between households and schools [16], the serial intervals of the Delta and Omicron variants [17], and the transmission depending on the vaccination status of infector and infectee [18].

In this study we aim to assess whether these routinely collected transmission pairs can provide additional information on transmission processes that could complement other data sources, such as contact tracing data or household studies. For this we first established that the infectees with an identified infector were a representative subset of all reported cases. Second, we verified that the linking of cases yielded plausible estimates for epidemiological characteristics, by comparing them to literature. With these two requirements met, we investigated the frequency of transmission events within and between groups of cases stratified by age group, transmission setting and vaccination status. Identification of characteristics that are associated with onward transmission within and between groups can suggest possible interventions to reduce transmission in the population.

## Methods

### Data

All laboratory-confirmed SARS-CoV-2 infections in the Netherlands were centrally registered in the OSIRIS database [14] using a unique identification number. These reported COVID-19 cases were coupled to their demographic characteristics, such as age, sex and postal code of the place of residence, as registered in the Personal Records Database [19]. Epidemiological characteristics, such as symptom onset date and transmission setting, were ascertained and registered by the public health officer. The public health officer would also ask the case by whom they thought they were infected. If this person could be identified and if this person was also present in the database as a previously reported case, its unique identification number was stored in the reported case file as “most probable infector”. By linking the identification numbers of the reported case and its most probable infector, transmission pairs were constructed with known characteristics for both infector and infectee. These characteristics include age, sex, postal code, residential province, date of report, date of symptom onset, date of testing positive for SARS-CoV-2 (i.e. date of laboratory confirmation), vaccination status and transmission setting.

We included data in the period from 27 February 2020 when the first case was reported until 11 April 2022, after which date a positive self test did not need to be confirmed anymore by the public health service [20] and the number of reported cases dropped. We completely excluded impossible transmission chains, such as self infections (of type A→A), reciprocal infections (of type A→B→A) and triangular infection chains (of type as A→B→C→A). The dates of testing positive were generally provided by automated systems of testing laboratories, whereas the dates of symptom onset were entered manually. Therefore we deemed dates of testing positive to be more trustworthy than symptom onset dates. For this reason, transmission pairs with an unrealistic period between testing positive were excluded from the data set, while pairs with an unrealistic period between symptom onset were only excluded from the serial interval analysis. We set these unrealistic periods between testing positive and between symptom onsets to be less than -10 days or more than 30 days, which is a bit broader than the reported serial interval distributions of smaller data sets [1,21].

The transmission setting (of the infectee) could be one or more settings from a list of options. They were assigned a single setting using the hierarchy: “Household”, “School”, “Work”, “Visit”, or “Other”. An exception was made for cases infected at nursing homes or long-term care facilities, who were infected at home, but were assigned the setting “Other”. The postal codes of infector and infectee were used to correct the settings in two ways. First, infectees who reported to have been infected at home by an infector with a postal code that differed from the infectee’s postal code, were assigned a “Visit” setting. Second, transmission pairs with unknown setting but with identical postal codes were classified as having a “Household” setting. It should be noted here that when an infector and infectee had identical postal codes they were not necessarily household members, but a validation study showed they were in 97% of the cases [22]. When using the term “Household” setting throughout the manuscript, it should be kept in mind that some of these transmission pairs might have occurred between neighbours.

### Analysis

To evaluate whether the reported cases with identified infector are representative of all reported cases, we calculated the number of infectees as a fraction of the total number of reported cases [23], stratified by 10-year age group, sex, residential province, calendar week, and whether the symptom onset date is known. Comparable fractions in the strata imply comparable representation. We also calculated the post-stratification weights. Post-stratification weights above one imply underrepresentation of the transmission pairs in these strata, and vice versa.

To check whether the transmission pairs were plausible, we compared the serial interval to values reported in literature [24]. The serial interval is defined as the time between symptom onset dates of infector and infectee. We calculated the mean and standard deviation of the serial intervals from all pairs with known symptom onset dates. For a comparison to values reported in literature, we calculated the mean serial interval by period with different dominant SARS-CoV-2 variants. The symptom onset date of the infector was used to assign each transmission pair to a period where the ancestral lineage (until 14 Feb 2021), the Alpha variant ( 15 Feb 2021 - 27 Jun 2021), the Delta variant (28 Jun 2021 - 26 Dec 2021) or the Omicron variant (27 Dec 2021 onwards) constituted the majority of tested samples [25].

To demonstrate how the transmission pair data can provide insights in transmission, we examined the transmission pairs by age groups of infectors and infectees. We first made a distinction by transmission setting, plotting the numbers of transmission pairs by 5-year age groups (up to 95+ years of age) and setting over the full study period. Next, we stratified the transmission pairs by vaccination status of infector and infectee, in 10-year age groups (up to 70+ years of age) to align with available vaccination coverage data [26,27]. We used data of infectees testing positive in September 2021 when the Delta variant was dominant. Before this month all persons of 12 years and older had the chance to get vaccinated, and after this month the effect of vaccine waning and the autumn booster campaign may impact the results. Persons who finished their vaccination series (i.e. receiving their second mRNA vaccine dose or their first vector vaccine dose) at least two weeks before testing positive were considered to be fully vaccinated [28,29]; all others - including partially vaccinated persons - were classified as not (fully) vaccinated. We note that the proportion of partially vaccinated persons was relatively small in September 2021 as the vaccination campaign was nearing its end [26]. We tested the number of transmission pairs in each combination of infector and infectee age group against the hypothesis that vaccination did not affect infection nor transmission and that mixing is independent of vaccination status, using a likelihood ratio test [30]. In this test, we compared the observed number of transmission pairs to the expected number under the null hypothesis based on the vaccination coverage in the general population (see Suppl S1). As a sensitivity analysis we repeated the analysis for the longer autumn period Sep - Nov 2021.

## Results

On 11 April 2022, in total 8,003,008 test-positive COVID-19 cases were reported in the OSIRIS database. For 713,222 of these cases, the most probable infector was identified. After removing 11,286 pairs with implausible intervals between dates of testing positive, 5,178 self infections, 18,204 reciprocal infections, and 72 triangular infection chains, the data consisted of 678,482 plausible transmission pairs, i.e., 8.5% of all reported cases. The transmission setting of 54,029 pairs (8% of all pairs) reported at home but between persons with a different postal codes were classified as “Visit”, and 24,714 pairs (3.6% of all pairs) with a missing transmission setting between persons with an identical postal code were assigned a “Household” setting. We assessed these pairs by evaluating their representativeness, serial interval and transmission patterns.

### Representativeness of transmission pairs

Stratifying the 678,482 infectees of the transmission pairs by age group, we found that all age groups are similarly represented, with post-stratification weights between 0.80 for 50-69 year olds and 1.21 for 30-39 year olds (Tab. 1). Infectees were proportionally divided over the sexes (Tab. 1), also within age groups (Fig. 1A). There were only 6 infectees with a missing value for age and 18 with a missing value for sex. The distribution of cases with identified infector over the Dutch provinces reasonably corresponded to that of all reported cases (Tab. 1 and Fig. 1B).

**Figure 1:**
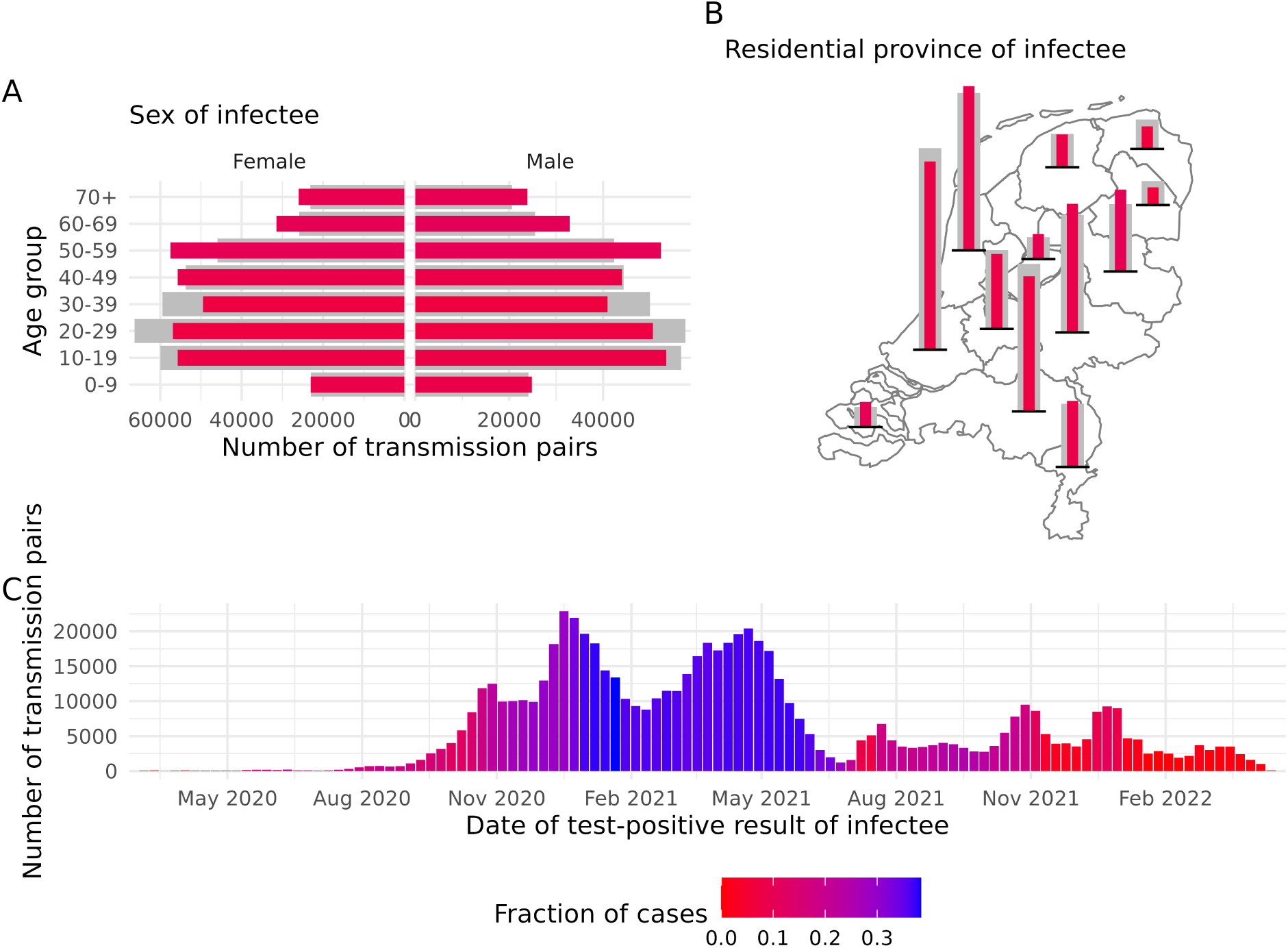
Number of transmission pairs during the period 27 February 2020 - 11 April 2022, stratified by A) sex and age group, B) residential province, and C) date of testpositive result of infectee, and coloured by the percentage of the total number of reported cases. The number of transmission pairs multiplied by the post-stratification weights are shown in grey bars for sex and age group (panel A) and residential province (panel B), indicating the extent of under- or overrepresentation. Note the colour legend applies to all subplots: the similar colours in panels A and B show better representativeness than the diverging colours in panel C.

**Table 1:** Overview of transmission pairs, stratified by age group, sex, residential province, transmission setting and known symptom onset date. The total number of cases and the number of cases with an identified infector in each stratification are shown for the period 27 February 2020 - 11 April 2022. The last columns give the percentage of cases with an identified infector and the post-stratification weight.

|  |  | Total number of reported cases | Number of cases with identified infector | % | Post-stratification weight |
|---|---|---|---|---|---|
| Age group | 0-9 | 557,769 | 47,857 | 8.6 | 0.98 |
|  | 10-19 | 1,380,645 | 109,123 | 7.9 | 1.07 |
|  | 20-29 | 1,466,960 | 107,445 | 7.3 | 1.15 |
|  | 30-39 | 1,296,606 | 90,420 | 7.0 | 1.21 |
|  | 40-49 | 1,161,847 | 99,728 | 8.6 | 0.98 |
|  | 50-59 | 1,044,714 | 109,760 | 10.5 | 0.80 |
|  | 60-69 | 607,691 | 64,298 | 10.6 | 0.80 |
|  | 70+ | 517,529 | 49,845 | 9.6 | 0.88 |
|  | (Missing) | 655 | 6 | 0.9 |  |
| Sex | Female | 4,227,660 | 355,689 | 8.4 | 1.00 |
|  | Male | 3,798,330 | 322,775 | 8.5 | 0.99 |
|  | (Missing) | 8,426 | 18 | 0.2 |  |
| Residential province | Drenthe | 201,457 | 12,407 | 6.2 | 1.37 |
|  | Flevoland | 182,709 | 17,592 | 9.6 | 0.88 |
|  | Fryslân | 276,611 | 23,004 | 8.3 | 1.02 |
|  | Gelderland | 980,605 | 90,576 | 9.2 | 0.91 |
|  | Groningen | 247,081 | 15,907 | 6.4 | 1.31 |
|  | Limburg | 527,012 | 46,654 | 8.9 | 0.95 |
|  | Noord-Brabant | 1,231,261 | 95,417 | 7.7 | 1.09 |
|  | Noord-Holland | 1,313,880 | 115,846 | 8.8 | 0.96 |
|  | Overijssel | 563,015 | 57,691 | 10.2 | 0.82 |
|  | Utrecht | 660,110 | 52,857 | 8.0 | 1.05 |
|  | Zeeland | 167,415 | 17,825 | 10.6 | 0.79 |
|  | Zuid-Holland | 1,683,205 | 132,706 | 7.9 | 1.07 |
|  | (Missing) | 55 | 0 | 0.0 |  |
| Transmission setting | Household | * | 412,011 |  |  |
|  | School | * | 18,008 |  |  |
|  | Work | * | 37,984 |  |  |
|  | Visit | * | 173,363 |  |  |
|  | Other | * | 25,317 |  |  |
|  | (Missing) | * | 11,799 |  |  |
| Symptom onset | Unknown | 4,555,121 | 82,723 | 1.8 | 4.65 |
|  | Known | 3,479,295 | 595,759 | 17.1 | 0.49 |

\* The transmission settings are only given for the number of cases with identified infector, as they were corrected with the use of postal code information and cannot be compared with the transmission settings of all reported cases.

The vast majority (86.3%) of transmission pairs was reported in the household or during visits (Tab. 1), which were settings where infector and infectee were well acquainted. The symptom onset date of cases with identified infector was more often known than for all reported cases (Tab. 1). The identification of infectors fluctuated heavily over time (Fig. 1C). Identification percentages were low at the start of the pandemic when testing for SARS-CoV-2 was not available for the general population and at the end of the pandemic when case numbers were high during the Omicron waves in the Netherlands. The percentages of identified infectors were highest during early 2021 when the Alpha variant emerged.

### Serial interval of transmission pairs

For 80% of the transmission pairs the symptom onset date was known for both infector and infectee. For each of these pairs, we calculate the time between these symptom onset dates, i.e., the serial interval. Of all serial intervals 666 (0.1%) were was discarded because they were lower than -10 days or longer than 30 days. The serial interval distribution peaked at 3 days (Fig. 2), with a mean of 3.6 days, a median of 3 days, and a standard deviation of 3.4 days. A fraction of 5% of the serial intervals was negative, indicating presymptomatic transmission. The mean serial interval was 3.7 days (sd 3.6 days) in the period dominated by the ancestral lineage, 3.6 days (sd 3.3 days) in the Alpha-dominated period, 3.6 days (sd 3.1 days) in the Delta-dominated period, and 3.4 days (sd 3.2 days) in the Omicron-dominated period.

**Figure 2:**
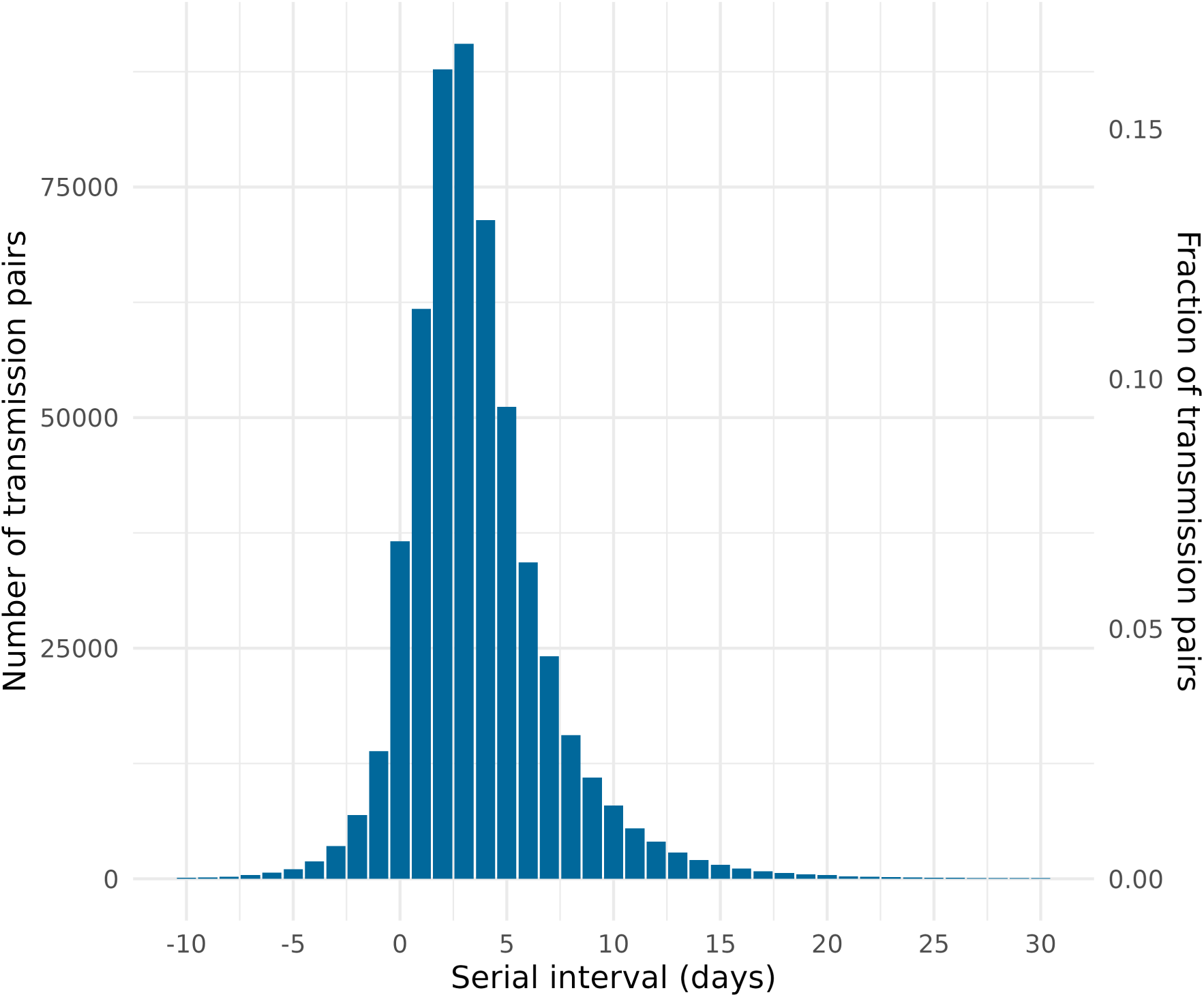
Serial interval distribution of transmission pairs where symptom onset dates of both infectee and infector are known, collected in the period 27 February 2020 - 11 April 2022. Only values from -10 days up to and including 30 days are deemed realistic.

### Transmission patterns

The transmission pair data allowed us to study transmission between groups with different characteristics. As an illustration, we evaluate transmission patterns between different age groups, stratified by setting or vaccination status.

Household transmissions showed a clear pattern along the diagonal and two off-diagonal parallels when depicted as a transmission matrix by 5-year age groups of infectors and infectees in households (Fig. 3). These household transmissions occurred between partners, flatmates or siblings (diagonal), from parent to child (upper parallel), or from child to parent (lower parallel). The transmission pattern in the “Visit” setting contained mainly transmissions between 15-24 year olds, but also showed intergenerational transmissions. Compared to the “Household” setting, these were shifted to higher ages where children and parents do not generally share a household. In the “School” setting many transmissions between children were reported. School transmissions from adults to children were more common than from children to adults, but it should be noted that an adult infected at school could have reported this with a “Work” setting. Indeed, in the “Work” setting some child to adult transmissions are reported. The “Work” setting was the only setting where transmissions occurred reasonably homogeneously between working age groups.

**Figure 3:**
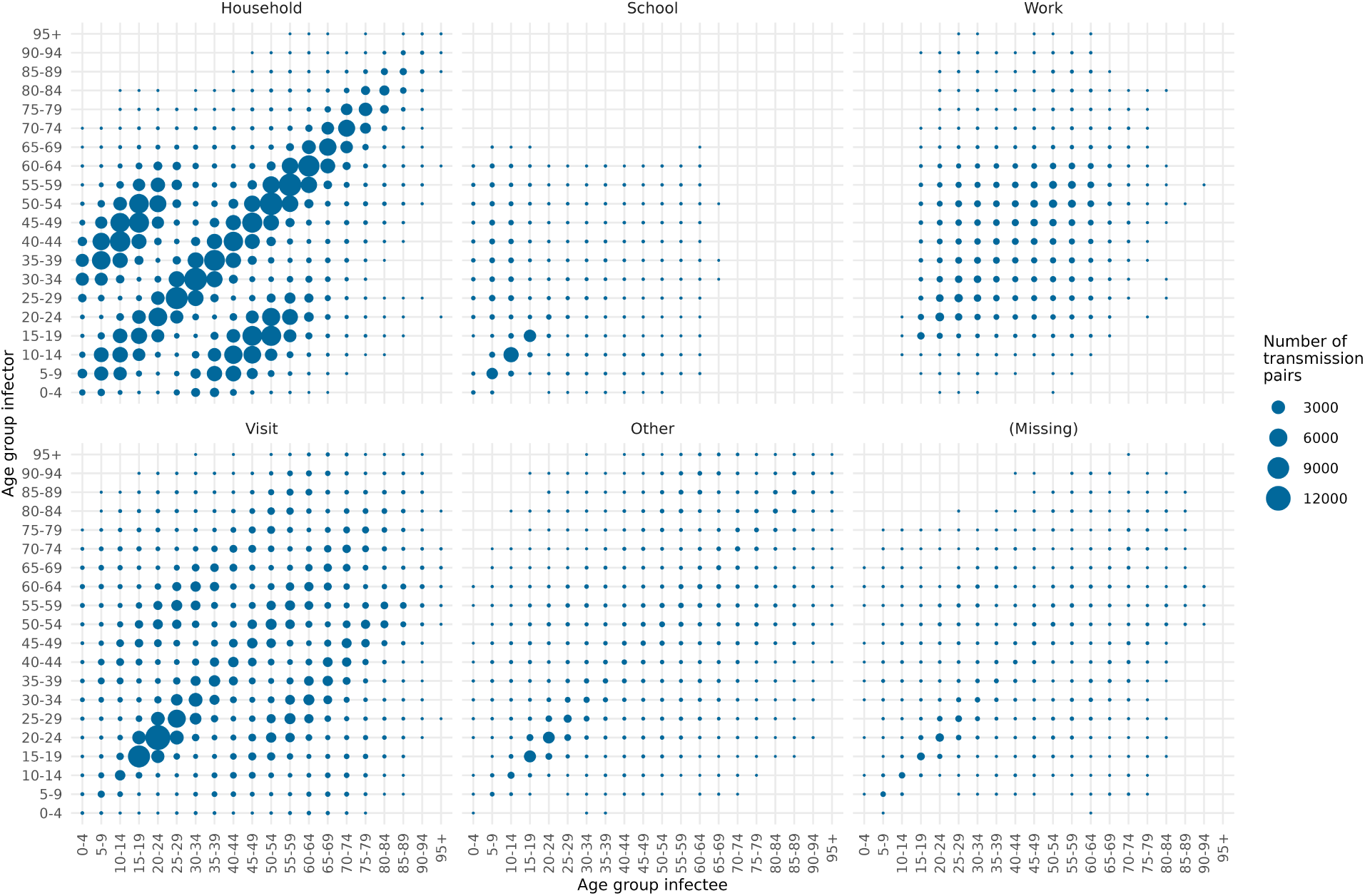
Transmission pairs stratified by transmission setting, between 5-year age groups of infectors and infectees, collected in the period 27 February 2020 - 11 April 2022. The size of the circles denotes the number of transmission pairs. Combinations of transmission setting, infector age group and infectee age group that contain less than 5 transmission pairs are not shown.

Stratifying the transmission pairs by vaccination status revealed a distinct transmission pattern, as illustrated here for September 2021 with in total 13,072 transmission pairs (Fig. 4). In 52 of the 64 age group combinations, the number of transmission pairs stratified by vaccination status differed significantly (p value < 0.001) from the number that would be expected if vaccination would not have any effect and if mixing were independent of vaccination status (see Tab. S1 in Supplement for values). Most transmission pairs were reported between persons who are not (fully) vaccinated, and the numbers were always larger than the expectation, as shown by the positive residual deviances (in red) in the lower left quadrant of Fig. 4. In the opposite quadrant transmissions between fully vaccinated persons were less common (in blue) than would be expected from the vaccination coverage alone.

**Figure 4:**
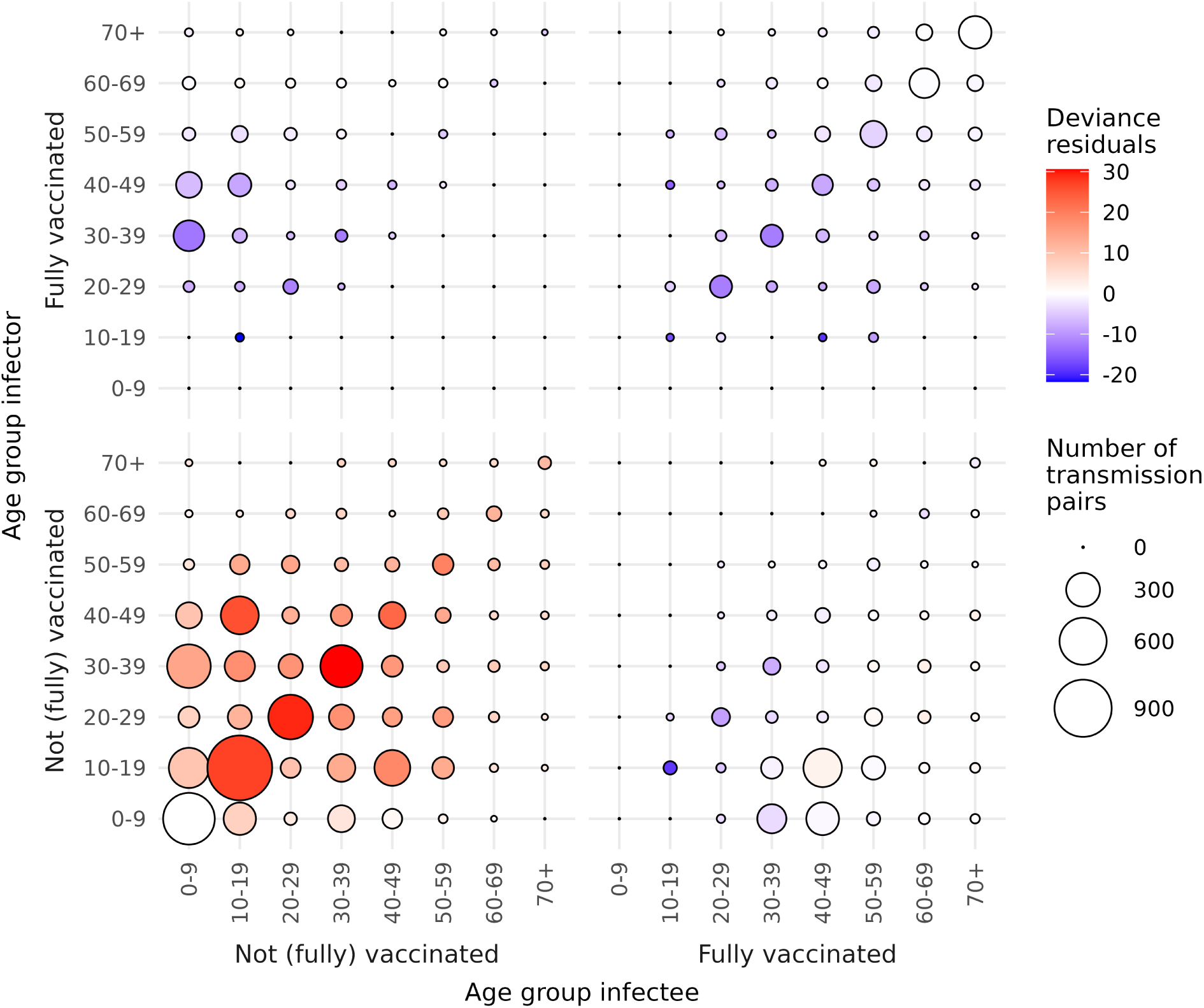
Transmission pairs stratified by vaccination status, between 10-year age groups of infectors and infectees, in September 2021. The size of the circles denotes the number of transmission pairs, coloured by the deviance residuals of the likelihood ratio test whether the number of transmission pairs is independent of the vaccination status. Red (blue) colours denote that the number of transmission pairs in the age group combination was higher (lower) than would be expected from the vaccination coverage alone. Combinations of vaccination status and age groups that contain less than 5 transmission pairs are not shown.

The results in both quadrants could be explained by the effect of vaccination, but this could be modified to an extent by assortative mixing [31]. Of all transmission pairs, 76% occur between persons of identical vaccination status, which indeed suggests assortative mixing. This could partly explain why the observed number of transmission pairs is higher than the expectation among not (fully) vaccinated persons. Similarly, the lower than expected numbers in the lower right and upper left quadrants could be partly explained by persons preferring not to mix with persons of opposing vaccination status. A notable exception in the lower right quadrant is that transmissions on the lower off-diagonal occurred more often than expected. These were most likely household transmissions from not (fully) vaccinated children to fully vaccinated parents, suggestive of a “leaky” rather than “all or nothing” protection by the vaccine.

An interesting group to consider are the 0-9 year olds because they were all unvaccinated. This age group more often infects not (fully) vaccinated than fully vaccinated other age groups (bottom row of Fig. 4), indicating vaccination protects against infection. In the other direction an even larger discrepancy shows that 0-9 year olds are more often infected by not (fully) vaccinated than fully vaccinated other age groups (left column of Fig. 4). This would suggest that vaccination has an additional effect on onward transmission. Assortativity may also play a role here but would probably not be the only explanation for the observed effects.

As a sensitivity analysis we repeated the analysis over a longer period including September, October and November 2021 with in total 56,206 transmission pairs. The general transmission pattern (see Fig. S1) is similar to the pattern shown in Fig. 4 with one striking difference: the number of observed transmission pairs among fully vaccinated persons in the oldest age groups is here higher than expected. This could be explained by a waning vaccine effect, as the oldest age groups were vaccinated first. Another explanation could be the assortative mixing effect, because also among the not (fully) vaccinated persons in the oldest age groups, the observations are higher than expected. A combination of both explanations is likely.

## Discussion

Routine surveillance of COVID-19 cases in the Netherlands allowed identifying a large number of transmission pairs, where the reported case could be linked to its most probable infector. The infectees of the transmission pairs were largely representative of all reported cases with regard to 10-year age group, sex and residential province. Because of this representativeness and because the transmission pairs were likely real transmission events (evidenced by the plausible serial interval distribution), we could use the observed transmission pairs as a proxy for all transmissions to study transmission patterns.

The transmission pair data were not representative for transmission events to reported cases over time. The fraction of cases with identified infector was low in some periods, especially during the first wave at the start of 2020 and after the emergence of Omicron at the end of 2021. The largest number of transmission pairs was collected during the period when the ancestral lineage or the Alpha variant were dominant. The household and visit transmission settings were likely overrepresented, because in these settings infectees could most probably identify their infector. On the other hand, transmissions from and to asymptomatic cases and other underreported cases are largely absent. These limitations are however also present in contact tracing data.

As plausibility check of the transmission pair data, we found a serial interval with an overall mean of 3.6 days, composed of mean serial intervals of 3.7 days, 3.6 days, 3.6 days and 3.4 days for the ancestral lineage, Alpha, Delta and Omicron variants respectively. These values were in line with a meta analysis of reported values [24] that found a pooled mean serial interval of 4.8 days (range of reported means 1.5 - 7.5 days), 3.5 days (2.4 - 4.5 days), 3.6 days (2.3 - 6.7 days), and 3.2 days (2.4 - 4.2 days) for the ancestral lineage, Alpha, Delta and Omicron BA.1 variants respectively. At the start of the pandemic long serial intervals were reported [2,32], but after the first pandemic peak when interventions were put in place the serial interval was found to be shorter [33–35]. Most of our transmission pairs were observed when interventions were in effect, explaining the relatively short serial interval for the ancestral lineage. We found 5% of the serial intervals to be negative. Other studies report higher negative fractions between 5.8% and 13% [1,12,33,36] or no negative fractions [2,32]. Negative serial intervals are an indication of presymptomatic transmission as was demonstrated for COVID-19 [37,38].

As the transmission pairs were largely self-reported and unconfirmed by genomic sequencing or field epidemiological investigation, they could have been misclassified in two ways. First, the identified infector was not necessarily the true infector. For confirmation additional testing of infector and infectee is required. In a previous study [17], we studied a period of two weeks with cocirculation of the Delta and Omicron BA.1 variants, that could be distinguished by S gene target failure (SGTF). Of the 1143 transmission pairs where both infector and infectee were tested, 27 pairs (2.4%) had conflicting results. This fraction is a lower bound to the true misclassification of infectors. Second, the direction of transmission could have been misclassified, where the identified infector was in fact the infectee. This could occur when infectees have a tendency to identify an infector with an earlier symptom onset. Such a tendency would lead to a discontinuity in the serial interval distribution at a serial interval of 0 days. As we did not observe such a discontinuity in our data (Fig. 2), we conclude there is no systemic misclassification in the transmission direction.

The added value of the transmission pair data lies in the adequate representativeness and the large size of the data set, which can reveal general transmission patterns. This was illustrated by studying transmissions between different age groups:

- Age-specific patterns in transmission pairs stratified by setting (Fig. 3) are similar to age-specific patterns in self-reported contacts by setting [39,40]. The similarity provides evidence favouring the ‘social contact hypothesis’ that proposes conversational contacts as a proxy for events at risk of transmission [41].
- Transmission pairs stratified by vaccination status compared to the vaccination coverage (Fig. 4) reveal that not (fully) vaccinated persons were more often infected than expected, which can be explained by a combination of the effect of vaccination and assortative mixing of persons with identical vaccination status.
- Fully vaccinated persons were less likely to be infected by the completely unvaccinated youngest age group, suggesting that the vaccine was effective against infection. Once infected, fully vaccinated persons were less likely to infect the youngest age group, suggesting that the vaccine was effective against onward transmission. These two types of vaccine effectiveness have been demonstrated in household studies [42], but the transmission pair data provided evidence from other settings and in a higher resolution.
- Our data suggested that persons preferably mix with persons who had an identical vaccination status, which has also been observed [43,44]. This assortativity introduces heterogeneity in the population structure that reduces the overall impact of vaccination [43,45].

The effects of vaccination (on infection and onward transmission) and assortativity could be estimated simultaneously [31], although it should be reminded that the not (fully) vaccinated population consists of unvaccinated and partially vaccinated persons. This would most likely underestimate both the effects of vaccination and assortativity.

Of all reported cases 8.5% could be linked to a most probable infector. This percentage was even higher than 30% from December 2020 till June 2021 during a crucial period of the pandemic when the Alpha variant emerged and large-scale vaccination was rolled out. A reported case would lack an infector when they did not know by whom they were infected (even if they did know the time and setting), when the case was not asked by or did not reply to the public health services for follow-up questions, or when the identified infector was not a reported case itself. This latter requisite for the linking of a transmission pair sets this system apart from contact tracing and has implications for its applicability. First, in order to improve chances to find a transmission pair, all cases should be centrally reported. This only applies to notifiable diseases. Second, the symptomatic rate should be considerable. Even if a reported case were able to identify an asymptomatic infector, asymptomatic cases are nearly always unreported and would not be in the data. Third, the number of cases should be (or at least have the potential to become) considerable. When the number of cases is small, most transmission links will be found through contact tracing. When the number of cases increases, the required contact tracing may exceed capacity, at which point the linking of transmission pairs from a database of reported cases can complement the data or provide a cheaper alternative. For these reasons this system would be most suited for (re)emerging pathogens with pandemic potential.

Transmission pair data can be used for a range of other research topics. The serial interval distribution can inform models to estimate the generation time distribution and the proportion of presymptomatic infections [37,46]. Building on the social contact hypothesis, the transmission pair data can be combined with contact data to infer age-specific epidemiological characteristics. The relative susceptibility of infectees and the relative infectiousness of infectors can be estimated simultaneously because the data contains the direction of transmission [47]. Also the spatial information of the transmission pairs could be a valuable data source, for instance, to study whether non-pharmaceutical interventions affect the distance between non-household transmission pairs. Aggregating the transmission pair data by infector yields the offspring distribution of the number of secondary infections per infector, which can be used to study superspreading potential [48,49] or to provide parametrisation of network models.

## Conclusions

Collecting transmission pair data is a valuable addition to routine surveillance. By identifying and linking infectors and infectees, detailed information can be generated about infection characteristics and transmission patterns of (re)emerging pathogens with pandemic potential. Although the implementation requires some adaptation of registration systems and some additional effort from public health officers, the resulting data set is an important resource for understanding how diseases spread. Furthermore, engaging citizens in the identification process can raise public awareness about transmission routes and prevention measures, strengthening community involvement in disease control [50]. Most importantly, timely collection and analysis of transmission pair data can provide crucial evidence to guide public health policy and interventions. Establishing such procedures now will enhance preparedness and response capacity for future pandemics, making it a worthwhile investment for public health systems.

## Data Availability

All data produced are available online at Zenodo

https://zenodo.org/record/18269559

## Declarations

### Ethics approval and consent to participate

The RIVM Centre for Clinical Expertise verified whether this study complies with the Dutch law for Medical Research Involving Human Subjects (WMO). In their opinion a review by an ethical research committee or institutional review board is not necessary by current national and European legislation (study number EPI-791). The use of data in our study is covered by the surveillance tasks of the RIVM, as defined in the Law on the RIVM (Article 3). Under these tasks, the RIVM is permitted to receive pseudonymised data without obtaining individual consent for each case, in accordance with the Public Health Act, Article 6c (Wet Publieke Gezondheid, artikel 6c). This research was conducted in accordance with the Declaration of Helsinki.

### Consent for publication

Not applicable

### Availability of data and materials

The datasets generated and/or analysed during the current study are available in the Zenodo repository (https://zenodo.org/record/18269559). The code used for data cleaning and the analyses is shared on github (https://github.com/rivm-syso/transmission-pairs-description).

## Competing interests

The authors declare that they have no competing interests.

## Funding

This study was funded by the Ministry of Health, Welfare and Sport (VWS) in the Netherlands.

## Authors’ contributions

JB designed the analysis, analyzed the data, and wrote the first draft of the manuscript. KYL and SA analyzed the data and reviewed the code. JvdK designed the first analyses and interpretations. IV contributed to data acquisition. SH contributed to the conception and implementation of the new surveillance tool. JW contributed to the conception of the new surveillance tool, and the interpretation of results. All authors contributed in writing the manuscript, and they read and approved the final manuscript.

## Acknowledgements

The authors would like to thank Pauline Bakker and Thomas Otten for critically assessing the manuscript.

## Notes

### Competing Interest Statement

The authors have declared no competing interest.

### Author Declarations

The RIVM Centre for Clinical Expertise verified whether this study complies with the Dutch law for Medical Research Involving Human Subjects (WMO). In their opinion a review by an ethical research committee or institutional review board is not necessary by current national and European legislation (study number EPI-791). The use of data in our study is covered by the surveillance tasks of the RIVM, as defined in the Law on the RIVM (Article 3). Under these tasks, the RIVM is permitted to receive pseudonymised data without obtaining individual consent for each case, in accordance with the Public Health Act, Article 6c (Wet Publieke Gezondheid, artikel 6c).

### Summary of Updates

Analysis of transmission pattern stratfied by age group and vaccination status reinterpreted Sensitivity analysis added Figure 4 corrected Results and discussion rewritten

## References

1. Du Z, Xu X, Wu Y, Wang L, Cowling BJ, Meyers LA. Serial Interval of COVID-19 among Publicly Reported Confirmed Cases. Emerging Infectious Diseases. 2020;26: 1341–1343. doi:10.3201/eid2606.200357

2. Liu T, Qi L, Yao M, Tian K, Lin M, Jiang H, et al. Serial Interval and Reproductive Number of COVID-19 Among 116 Infector-infectee Pairs - Jingzhou City, Hubei Province, China, 2020. China CDC weekly. 2020;2: 491–495. doi:10.46234/ccdcw2020.118

3. Xu X-K, Liu XF, Wu Y, Ali ST, Du Z, Bosetti P, et al. Reconstruction of Transmission Pairs for Novel Coronavirus Disease 2019 (COVID-19) in Mainland China: Estimation of Superspreading Events, Serial Interval, and Hazard of Infection. Clinical Infectious Diseases: An Official Publication of the Infectious Diseases Society of America. 2020;71: 3163–3167. doi:10.1093/cid/ciaa790

4. Sun K, Wang W, Gao L, Wang Y, Luo K, Ren L, et al. Transmission heterogeneities, kinetics, and controllability of SARS-CoV-2. Science. 2021;371: eabe2424. doi:10.1126/science.abe2424

5. Miura F, Backer JA, Rijckevorsel G van, Bavalia R, Raven S, Petrignani M, et al. Time Scales of Human Mpox Transmission in The Netherlands. The Journal of Infectious Diseases. 2024;229: 800–804. doi:10.1093/infdis/jiad091

6. Cauchemez S, Carrat F, Viboud C, Valleron AJ, Boëlle PY. A Bayesian MCMC approach to study transmission of influenza: Application to household longitudinal data. Statistics in Medicine. 2004;23: 3469–3487. doi:10.1002/sim.1912

7. Zhang C, Fang VJ, Chan K-H, Leung GM, Ip DKM, Peiris JSM, et al. Interplay Between Viral Shedding, Age, and Symptoms in Individual Infectiousness of Influenza Cases in Households. The Journal of Infectious Diseases. 2025;231: 462–470. doi:10.1093/infdis/jiae434

8. Li F, Li Y-Y, Liu M-J, Fang L-Q, Dean NE, Wong GWK, et al. Household transmission of SARS-CoV-2 and risk factors for susceptibility and infectivity in Wuhan: A retrospective observational study. The Lancet Infectious Diseases. 2021;21: 617–628. doi:10.1016/S1473-3099(20)30981-6

9. Dattner I, Goldberg Y, Katriel G, Yaari R, Gal N, Miron Y, et al. The role of children in the spread of COVID-19: Using household data from Bnei Brak, Israel, to estimate the relative susceptibility and infectivity of children. PLOS Computational Biology. 2021;17: e1008559. doi:10.1371/journal.pcbi.1008559

10. Reukers DFM, Boven M van, Meijer A, Rots N, Reusken C, Roof I, et al. High Infection Secondary Attack Rates of Severe Acute Respiratory Syndrome Coronavirus 2 in Dutch Households Revealed by Dense Sampling. Clinical Infectious Diseases: An Official Publication of the Infectious Diseases Society of America. 2022;74: 52–58. doi:10.1093/cid/ciab237

11. Bistaraki A, Roussos, Tsiodras, Sypsa V and. Age-dependent effects on infectivity and susceptibility to SARS-CoV-2 infection: Results from nationwide contact tracing data in Greece. Infectious Diseases. 2022;54: 186–195. doi:10.1080/23744235.2021.1995627

12. Hong K, Yum S, Kim J, Chun BC. The Serial Interval of COVID-19 in Korea: 1,567 Pairs of Symptomatic Cases from Contact Tracing. Journal of Korean Medical Science. 2020;35: e435. doi:10.3346/jkms.2020.35.e435

13. Beest DE te, Wallinga J, Donker T, Boven M van. Estimating the generation interval of influenza A (H1N1) in a range of social settings. Epidemiology (Cambridge, Mass). 2013;24: 244–250. doi:10.1097/EDE.0b013e31827f50e8

14. Geubbels ELPE, Backer JA, Bakhshi-Raiez F, Beek RFHJ van der, Benthem BHB van, Boogaard J van den, et al. The daily updated Dutch national database on COVID-19 epidemiology, vaccination and sewage surveillance. Scientific Data. 2023;10: 469. doi:10.1038/s41597-023-02232-w

15. Hoek W van der, Backer JA, Bodewes R, Friesema I, Meijer A, Pijnacker R, et al. [The role of children in the transmission of SARS-CoV-2]. Nederlands Tijdschrift Voor Geneeskunde. 2020;164: D5140. Available: https://www.ntvg.nl/artikelen/de-rol-van-kinderen-de-transmissie-van-sars-cov-2

16. Van Iersel SCJL, Backer JA, Van Gaalen RD, Andeweg SP, Munday JD, Wallinga J, et al. Empirical evidence of transmission over a school-household network for SARS-CoV-2; exploration of transmission pairs stratified by primary and secondary school. Epidemics. 2023;43: 100675. doi:10.1016/j.epidem.2023.100675

17. Backer JA, Eggink D, Andeweg SP, Veldhuijzen IK, Maarseveen N van, Vermaas K, et al. Shorter serial intervals in SARS-CoV-2 cases with Omicron BA.1 variant compared with Delta variant, the Netherlands, 13 to 26 December 2021. Euro Surveillance: Bulletin Europeen Sur Les Maladies Transmissibles = European Communicable Disease Bulletin. 2022;27: 2200042. doi:10.2807/1560-7917.ES.2022.27.6.2200042

18. Van Dissel, Jaap. COVID-19 briefing tweede kamer, 21 dec 2021 (accessed June 2025). 2021. Available: https://www.tweedekamer.nl/sites/default/files/atoms/files/20211221_commissie_vws_briefing_presentatie_jaap_van_dissel.pdf

19. Government of the Netherlands. Personal Records Database (BRP). 2023. Available: https://www.government.nl/topics/personal-data/personal-records-database-brp

20. National Institute for Public Health and the Environment (RIVM). Tijdlijn van coronamaatregelen 2021 (Timeline of corona measures 2022). 2023. Available: https://www.rivm.nl/gedragsonderzoek/tijdlijn-maatregelen-covid-2022

21. Ryu S, Kim D, Lim J-S, Ali ST, Cowling BJ. Serial interval and transmission dynamics during SARS-CoV-2 Delta variant predominance, South Korea. Emerging Infectious Diseases. 2022;28: 507–510. doi:10.3201/eid2802.211472

22. Statistics Netherlands. Transmissieparen adres, werkgever en school, jan-sep 2021 [Transmission pairs address, employer and school, Jan-Sep 2021]. 2021. Available: https://www.cbs.nl/nl-nl/maatwerk/2021/49/transmissieparen-adres-werkgever-en-school-jan-sep-2021

23. National Institute for Public Health and the Environment (RIVM). Covid-19 karakteristieken per casus landelijk (Covid-19 case characateristics nationwide). 2023. Available: https://data.rivm.nl/meta/srv/dut/catalog.search#/metadata/2c4357c8-76e4-4662-9574-1deb8a73f724

24. Xu X, Wu Y, Kummer AG, Zhao Y, Hu Z, Wang Y, et al. Assessing changes in incubation period, serial interval, and generation time of SARS-CoV-2 variants of concern: A systematic review and meta-analysis. BMC Medicine. 2023;21: 374. doi:10.1186/s12916-023-03070-8

25. National Institute for Public Health and the Environment (RIVM). Varianten van het coronavirus SARS-CoV-2 (Variants of SARS-CoV-2 corona virus). 2026. Available: https://www.rivm.nl/corona/actueel/virusvarianten

26. Backer DAM Jantien A. AND Klinkenberg. Estimating the effectiveness of non-pharmaceutical interventions against COVID-19 transmission in the netherlands. PLOS Computational Biology. 2025;21: 1–17. doi:10.1371/journal.pcbi.1013502

27. Backer JA. Estimating the effectiveness of non-pharmaceutical interventions against COVID-19 transmission in the Netherlands. https://github.com/rivm-syso/effectiveness_NPIs; 2025. doi:10.21945/ba7f853e-749e-4d1c-8652-05c0b32261a3

28. Polack FP, Thomas SJ, Kitchin N, Absalon J, Gurtman A, Lockhart S, et al. Safety and efficacy of the BNT162b2 mRNA covid-19 vaccine. New England Journal of Medicine. 2020;383: 2603–2615. doi:10.1056/NEJMoa2034577

29. Baden LR, El Sahly HM, Essink B, Kotloff K, Frey S, Novak R, et al. Efficacy and safety of the mRNA-1273 SARS-CoV-2 vaccine. New England Journal of Medicine. 2021;384: 403–416. doi:10.1056/NEJMoa2035389

30. McDonald JH. Handbook of Biological Statistics. 3rd ed. Baltimore, Maryland: Sparky House Publishing; 2014. Available: http://www.biostathandbook.com/gtestgof.html

31. Geismar C, White PJ, Cori A, Jombart T. Sorting out assortativity: When can we assess the contributions of different population groups to epidemic transmission? PLoS ONE. 2024;19: e0313037. doi:10.1371/journal.pone.0313037

32. Li Q, Guan X, Wu P, Wang X, Zhou L, Tong Y, et al. Early Transmission Dynamics in Wuhan, China, of Novel Coronavirus–Infected Pneumonia. New England Journal of Medicine. 2020;382: 1199–1207. doi:10.1056/NEJMoa2001316

33. Ali ST, Wang L, Lau EHY, Xu X-K, Du Z, Wu Y, et al. Serial interval of SARS-CoV-2 was shortened over time by nonpharmaceutical interventions. Science. 2020;369: 1106–1109. doi:10.1126/science.abc9004

34. Ali ST, Yeung A, Shan S, Wang L, Gao H, Du Z, et al. Serial Intervals and Case Isolation Delays for Coronavirus Disease 2019: A Systematic Review and Meta-Analysis. Clinical Infectious Diseases. 2022;74: 685–694. doi:10.1093/cid/ciab491

35. Chen D, Lau Y-C, Xu X-K, Wang L, Du Z, Tsang TK, et al. Inferring time-varying generation time, serial interval, and incubation period distributions for COVID-19. Nature Communications. 2022;13: 7727. doi:10.1038/s41467-022-35496-8

36. Haddad N, Clapham HE, Abou Naja H, Saleh M, Farah Z, Ghosn N, et al. Calculating the serial interval of SARS-CoV-2 in Lebanon using 2020 contact-tracing data. BMC Infectious Diseases. 2021;21: 1053. doi:10.1186/s12879-021-06761-w

37. Ferretti L, Wymant C, Kendall M, Zhao L, Nurtay A, Abeler-Dörner L, et al. Quantifying SARS-CoV-2 transmission suggests epidemic control with digital contact tracing. Science. 2020;368: eabb6936. doi:10.1126/science.abb6936

38. He X, Lau EHY, Wu P, Deng X, Wang J, Hao X, et al. Temporal dynamics in viral shedding and transmissibility of COVID-19. Nature Medicine. 2020;26: 672–675. doi:10.1038/s41591-020-0869-5

39. Mossong J, Hens N, Jit M, Beutels P, Auranen K, Mikolajczyk R, et al. Social Contacts and Mixing Patterns Relevant to the Spread of Infectious Diseases. PLOS Medicine. 2008;5: e74. doi:10.1371/journal.pmed.0050074

40. Prem K, Zandvoort K van, Klepac P, Eggo RM, Davies NG, Group C for the MM of IDCW, et al. Projecting contact matrices in 177 geographical regions: An update and comparison with empirical data for the COVID-19 era. PLOS Computational Biology. 2021;17: e1009098. doi:10.1371/journal.pcbi.1009098

41. Wallinga J, Teunis P, Kretzschmar M. Using data on social contacts to estimate age-specific transmission parameters for respiratory-spread infectious agents. American Journal of Epidemiology. 2006;164: 936–944. doi:10.1093/aje/kwj317

42. De Gier B, Andeweg S, Backer JA, RIVM COVID-19 surveillance and epidemiology team, Hahné SJ, Van Den Hof S, et al. Vaccine effectiveness against SARS-CoV-2 transmission to household contacts during dominance of Delta variant (B.1.617.2), the Netherlands, August to September 2021. Eurosurveillance. 2021;26. doi:10.2807/1560-7917.ES.2021.26.44.2100977

43. Barclay VC, Smieszek T, He J, Cao G, Rainey JJ, Gao H, et al. Positive network assortativity of influenza vaccination at a high school: Implications for outbreak risk and herd immunity. PLoS ONE. 2014;9: e87042. doi:10.1371/journal.pone.0087042

44. Hâncean M-G, Lerner J, Perc M, Molina JL. Assortative mixing of opinions about COVID-19 vaccination in personal networks. Scientific Reports. 2024;14: 3385. doi:10.1038/s41598-024-53825-3

45. Fisman DN, Amoako A, Tuite AR. Impact of population mixing between vaccinated and unvaccinated subpopulations on infectious disease dynamics: Implications for SARS-CoV-2 transmission. CMAJ. 2022;194: E573–E580. doi:10.1503/cmaj.212105

46. Hart WS, Maini PK, Thompson RN. High infectiousness immediately before COVID-19 symptom onset highlights the importance of continued contact tracing. Elife. 2021;10. doi:10.7554/eLife.65534

47. Leung KY, Miura F, Backer JA. Disentangling infectiousness and susceptibility by age group using transmission pair data: A study of SARS-CoV-2 household transmission. medRxiv. 2026. doi:10.64898/2026.06.04.26354892

48. Endo A, Centre for the Mathematical Modelling of Infectious Diseases COVID-19 Working Group, Abbott S, Kucharski AJ, Funk S. Estimating the overdispersion in COVID-19 transmission using outbreak sizes outside china. Wellcome Open Res. 2020;5: 67. doi:10.12688/wellcomeopenres.15842.3

49. Kremer C, Torneri A, Boesmans S, Meuwissen H, Verdonschot S, Vanden Driessche K, et al. Quantifying superspreading for COVID-19 using poisson mixture distributions. Sci Rep. 2021;11: 14107. doi:10.1038/s41598-021-93578-x

50. Tan Y-R, Agrawal A, Matsoso MP, Katz R, Davis SLM, Winkler AS, et al. A call for citizen science in pandemic preparedness and response: Beyond data collection. BMJ Glob Health. 2022;7: e009389. doi:10.1136/bmjgh-2022-009389

