## Supplementary material for "Who infected the reported cases? Evidence from 678,482 COVID-19 cases with identified infector collected in routine surveillance in the Netherlands, 2020-2022"

* corresponding author

### S1 Likelihood ratio test for transmission matrix

For each combination of infectees of age group $i$ and infectors of age group $j$ we observe the total number of transmission pairs $N^{ij}$. These pairs are further stratified by vaccination status, which is either fully vaccinated $v$ or not (fully) vaccinated $u$, i.e. $N_{uu}^{ij}$, $N_{uv}^{ij}$, $N_{vu}^{ij}$, and $N_{vv}^{ij}$. Under the hypothesis that vaccination did not affect infection nor transmission and that mixing is independent of vaccination status, the expected number of transmission pairs in each stratum is calculated from the vaccination coverage $c_i$ and $c_j$ in age groups $i$ and $j$:

$$
\begin{aligned}
E_{uu}^{ij} &= (1 - c_i)(1 - c_j)N^{ij} \\
E_{uv}^{ij} &= (1 - c_i)c_j N^{ij} \\
E_{vu}^{ij} &= c_i(1 - c_j)N^{ij} \\
E_{vv}^{ij} &= c_i c_j N^{ij}
\end{aligned}
$$

To compare the goodness of fit we compute the likelihood ratio $\Lambda$ of the underlying multinomial model:

$$\Lambda_{kl}^{ij} = N_{kl}^{ij} \ln\left(\frac{N_{kl}^{ij}}{E_{kl}^{ij}}\right) - \left(N_{kl}^{ij} - E_{kl}^{ij}\right) \quad \forall\, \{kl\} \in \{uu, uv, vu, vv\}.$$

The G-statistic $G = 2\sum_{kl} \Lambda_{kl}^{ij}$ follows a $\chi^2$ distribution with 3 degrees of freedom. The signed deviance residual $r$ for each observation is computed with the sign function $\mathrm{sgn}()$:

$$r_{kl}^{ij} = \mathrm{sgn}(N_{kl}^{ij} - E_{kl}^{ij})\sqrt{2\Lambda_{kl}^{ij}} \quad \forall\, \{kl\} \in \{uu, uv, vu, vv\}.$$

Table 1: Results of likelihood ratio test to test whether the number of transmission pairs was independent of the vaccination status (fully vaccinated or not (fully) vaccinated), in each age group combination, in September 2021. The significance level is indicated by *** (p-value < 0.001), ** (p-value < 0.01) and * (p-value < 0.05).

| Infector age group | Infectee age group | Number of transmission pairs | p-value | |
|---|---|---|---|---|
| 0-9 | 0-9 | 728 | 1.00e+00 | |
| 0-9 | 10-19 | 274 | 6.54e-12 | *** |
| 0-9 | 20-29 | 47 | 2.92e-05 | *** |
| 0-9 | 30-39 | 403 | 5.02e-06 | *** |
| 0-9 | 40-49 | 375 | 6.17e-01 | |
| 0-9 | 50-59 | 55 | 1.21e-01 | |
| 0-9 | 60-69 | 30 | 9.31e-01 | |
| 0-9 | 70+ | 17 | 9.88e-01 | |
| 10-19 | 0-9 | 431 | 1.14e-18 | *** |
| 10-19 | 10-19 | 1,224 | 0.00e+00 | *** |
| 10-19 | 20-29 | 128 | 2.56e-29 | *** |
| 10-19 | 30-39 | 313 | 1.04e-38 | *** |
| 10-19 | 40-49 | 742 | 1.44e-151 | *** |
| 10-19 | 50-59 | 270 | 3.02e-57 | *** |
| 10-19 | 60-69 | 34 | 9.07e-05 | *** |
| 10-19 | 70+ | 25 | 1.48e-02 | * |
| 20-29 | 0-9 | 134 | 1.76e-23 | *** |
| 20-29 | 10-19 | 190 | 1.58e-52 | *** |
| 20-29 | 20-29 | 784 | 8.13e-263 | *** |
| 20-29 | 30-39 | 220 | 2.21e-93 | *** |
| 20-29 | 40-49 | 125 | 5.42e-63 | *** |
| 20-29 | 50-59 | 206 | 2.14e-66 | *** |
| 20-29 | 60-69 | 65 | 3.78e-18 | *** |

| Infector age group | Infectee age group | Number of transmission pairs | p-value | |
|---|---|---|---|---|
| 20-29 | 70+ | 22 | 4.77e-05 | *** |
| 30-39 | 0-9 | 757 | 2.11e-79 | *** |
| 30-39 | 10-19 | 282 | 4.21e-78 | *** |
| 30-39 | 20-29 | 192 | 1.36e-88 | *** |
| 30-39 | 30-39 | 699 | 7.99e-280 | *** |
| 30-39 | 40-49 | 183 | 2.22e-73 | *** |
| 30-39 | 50-59 | 68 | 3.29e-22 | *** |
| 30-39 | 60-69 | 77 | 4.80e-23 | *** |
| 30-39 | 70+ | 32 | 9.18e-13 | *** |
| 40-49 | 0-9 | 341 | 4.40e-28 | *** |
| 40-49 | 10-19 | 526 | 6.42e-205 | *** |
| 40-49 | 20-29 | 95 | 7.61e-47 | *** |
| 40-49 | 30-39 | 180 | 6.64e-78 | *** |
| 40-49 | 40-49 | 347 | 3.38e-140 | *** |
| 40-49 | 50-59 | 108 | 2.74e-48 | *** |
| 40-49 | 60-69 | 46 | 2.09e-09 | *** |
| 40-49 | 70+ | 49 | 9.14e-11 | *** |
| 50-59 | 0-9 | 59 | 1.17e-04 | *** |
| 50-59 | 10-19 | 164 | 2.62e-53 | *** |
| 50-59 | 20-29 | 141 | 4.83e-54 | *** |
| 50-59 | 30-39 | 73 | 4.35e-33 | *** |
| 50-59 | 40-49 | 108 | 4.55e-34 | *** |
| 50-59 | 50-59 | 325 | 5.87e-89 | *** |
| 50-59 | 60-69 | 88 | 1.16e-26 | *** |
| 50-59 | 70+ | 59 | 6.44e-14 | *** |
| 60-69 | 0-9 | 44 | 5.31e-01 | |

| Infector age group | Infectee age group | Number of transmission pairs | p-value | |
|---|---|---|---|---|
| 60-69 | 10-19 | 23 | 2.07e-02 | * |
| 60-69 | 20-29 | 42 | 4.90e-11 | *** |
| 60-69 | 30-39 | 60 | 2.62e-11 | *** |
| 60-69 | 40-49 | 32 | 2.51e-02 | * |
| 60-69 | 50-59 | 103 | 2.44e-18 | *** |
| 60-69 | 60-69 | 304 | 4.83e-39 | *** |
| 60-69 | 70+ | 80 | 3.81e-09 | *** |
| 70+ | 0-9 | 20 | 2.05e-03 | ** |
| 70+ | 10-19 | 7 | 5.90e-01 | |
| 70+ | 20-29 | 10 | 8.65e-01 | |
| 70+ | 30-39 | 18 | 1.75e-10 | *** |
| 70+ | 40-49 | 29 | 1.90e-10 | *** |
| 70+ | 50-59 | 47 | 2.50e-07 | *** |
| 70+ | 60-69 | 76 | 1.65e-08 | *** |
| 70+ | 70+ | 336 | 1.78e-32 | *** |

### S2 Sensitivity analysis

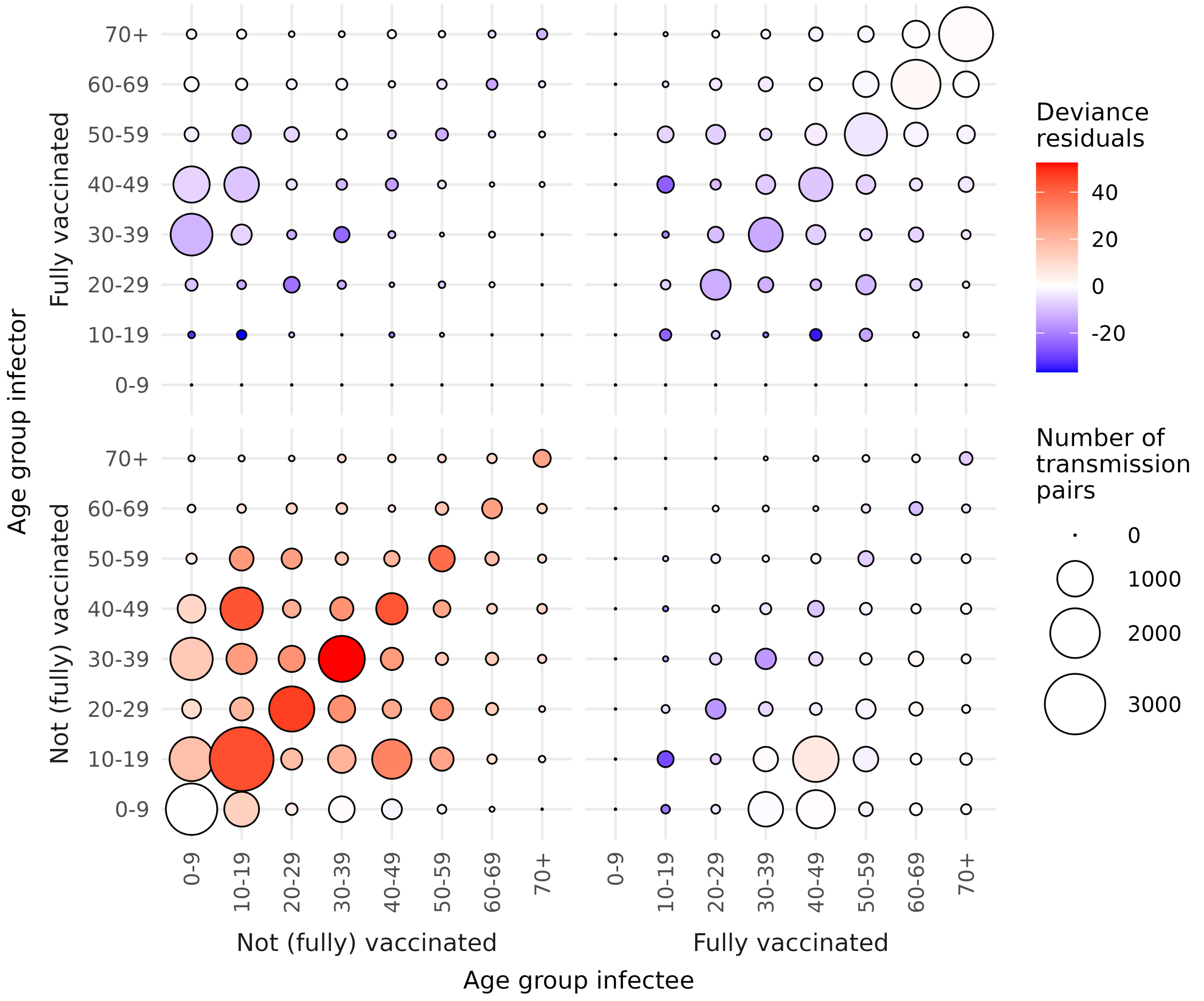


Figure 1: Transmission pairs stratified by vaccination status, between 10-year age groups of infectors and infectees, in the **period September-November 2021**. The size of the circles denotes the number of transmission pairs, coloured by the deviance residuals of the likelihood ratio test whether the number of transmission pairs is independent of the vaccination status. Red (blue) colours denote that the number of transmission pairs in the age group combination was higher (lower) than would be expected from the vaccination coverage alone. Combinations of vaccination status and age groups that contain less than 5 transmission pairs are not shown.
